# Challenges and opportunities of the COVID-19 pandemic for perinatal mental health care: a mixed methods study of mental health care staff

**DOI:** 10.1101/2020.09.23.20199927

**Authors:** CA Wilson, C Dalton-Locke, S Johnson, A Simpson, S Oram, LM Howard

## Abstract

**Purpose:** the aim of this study was to explore staff perceptions of the impact of the COVID-19 pandemic on mental health service delivery and outcomes for women who were pregnant or in the first year after birth (‘perinatal’ women).

**Methods:** secondary analysis of an online mixed-methods survey open to all mental health care staff in the UK involving 363 staff working with women in the perinatal period.

**Results:** staff perceived the mental health of perinatal women to be particularly vulnerable to the impact of stressors associated with the pandemic such as social isolation (rated by 79.3% as relevant or extremely relevant; 288/363) and domestic violence and abuse (53.3%; 192/360). As a result of changes to mental health and other health and social care services, staff reported feeling less able to assess women, particularly their relationship with their baby (43.3%; 90/208), and to mobilise safeguarding procedures (29.4%; 62/211). While 42% of staff reported that some women engaged poorly with virtual appointments, they also found flexible remote consulting to be beneficial for some women and helped time management due to reductions in travel time.

**Conclusions:** delivery of perinatal care needs to be tailored to the needs of women; virtual appointments are perceived not to be appropriate for assessments but may be helpful for some women in subsequent interactions. Safeguarding and other risk assessment procedures must remain robust in spite of modifications made to service delivery during pandemics.

## Introduction

Since the advent of the global COVID-19 pandemic in March 2020, numerous concerns regarding its impact on public mental health have been raised. These include the impact of lockdown and social distancing measures, bereavement and a range of other socio-economic stressors (Adhanom Ghebreyesus 2020). Surveys of adults from around the world have reported increased symptoms of anxiety and depression, with many finding women to be at an increased risk (González-Sanguino et al. 2020; Hyland et al. 2020; Pierce et al. 2020). There are other groups who are especially vulnerable to the adverse mental health impacts of the pandemic.

Women in the perinatal period (during pregnancy and up to one year postpartum) may be particularly in need of social support. However, having been advised at the peak of the pandemic to follow strict social distancing measures known in the UK as ‘shielding’, these women are experiencing reduced contact with friends, family and health and social care services (Caparros-Gonzalez and Alderdice 2020). Mental illness already affects at least one in five women in the perinatal period and is associated with adverse maternal and child outcomes (Ayers and Shakespeare 2015; Stein et al. 2014). There have been increases in violence, abuse and exploitation reported during the pandemic (Usher et al. 2020), also known to be associated with adverse maternal and child outcomes (Oram et al. 2017).

There are also concerns about the impact of the pandemic on those with existing mental illness (Sheridan Rains et al. 2020). Those with severe mental illness are at increased risk of COVID-19 infection due to higher rates of comorbidities such as physical ill health and socioeconomic adversities. They are also more vulnerable to the indirect effects of the pandemic such as reduced service provision and social isolation, which may precipitate relapse (Moreno et al. 2020).

There remains limited understanding of the impact of the pandemic on care provided for those with mental illness, including in the perinatal period (The Lancet Psychiatry 2020). However, there are reports of early discharges from inpatient psychiatric units and reduced community service provision, including fewer face-to-face meetings (Moreno et al. 2020). An online mixed-methods survey administered to staff working in mental health services in the UK sought to explore the impact on mental health care (Johnson et al. 2020). Here we report the responses from staff working with women in the perinatal period.

## Materials and methods

From 22 April 2020 to 12 May 2020, the NIHR Mental Health Policy Research Unit (MHPRU) surveyed staff working across services that provide mental health care in the UK, including the National Health Service (NHS), private services, social care and third sector or voluntary services. The development of the online questionnaire has been previously described and was done in close collaboration with service users and clinicians (Johnson et al. 2020). Both structured and open-ended questions explored staff’s perceptions of problems faced by service users and carers and challenges to care provision. These core questions were followed by additional questions specific to staff working in particular settings and specialities. Recruitment was via social media and relevant professional bodies, with a particular emphasis on recruiting a broad demographic, including staff with protected characteristics such as Black, Asian and Minority Ethnic groups. The King’s College London Research Ethics Committee approved the study (MRA-19/20-18372).

The sample comprised those who reported working with pregnant or postnatal women in generic or perinatal specialist services (community teams or inpatient Mother and Baby Units: MBUs) and who completed at least one question from each of the three main sections of questions open to all respondents described above, as per the approach taken by a previous analysis of the full sample of respondents (Johnson et al. 2020).

Quantitative data were analysed using Stata 15 (StataCorp 2007) and are presented as descriptive statistics. Qualitative data from open-ended questions were analysed using inductive semantic thematic analysis (Braun and Clarke 2006). Initial coding of data led to identification of potential themes, which were checked against coded extracts in a recursive process. For the purposes of providing context for the qualitative analysis, several of the authors are practising psychiatrists, some of whom work in perinatal psychiatry, although none were practising in perinatal services at the time of the pandemic. Peer debriefing was employed between CAW, who conducted the qualitative analysis, and LMH (Lincoln and Guba 1985).

## Results

### Participant characteristics

Of 2180 respondents who completed at least one question from each of the three main sections open to all respondents (3712 started the survey and 1793 reached the end of it), 363 reported working with perinatal service users in generic or specialist services. 56 of these reported only working in perinatal services. 85.2% of this sample of 363 staff described their gender as female and 70.3% their ethnic group as White British. The majority (91.2%) worked in the NHS and in England (82.4%).

Staff were asked what service delivery setting they currently work in; 255 worked in a community mental health team (CMHT). 54 reported working in a hospital inpatient service, 18 of these in MBUs. Other settings were crisis teams (N=78) (Johnson 2013) and community groups (N=26). 32.3% described their profession as a nurse, 13% as psychiatrist and 10.2% as psychologist. 15.8% identified as ‘other qualified therapist’. Other reported professions included social work and occupational therapy. 41.6% identified as a manager or lead clinician in their service.

### Difficulties experienced by users of perinatal mental health services

Staff were asked to rate the relevance, on a five-point scale, of pre-specified problems among service users and carers with whom they were currently in contact. Table 1 displays proportions of staff rating each item as ‘very relevant’ or ‘extremely relevant’; items are ordered by highest proportion first.

**Table 1:** difficulties rated as very or extremely relevant for mental health service users and their carers identified by mental health staff.

|  | n/N* | % |
| --- | --- | --- |
| Lack of access to usual support networks of family and friends | 288/363 | 79.3 |
| Loneliness due to or made worse by social distancing, self-isolation and/or shielding | 275/363 | 75.8 |
| Lack of usual work and activities | 245/358 | 68.4 |
| Worries about getting COVID-19 infection | 240/363 | 66.1 |
| Lack of access to usual support from other services (primary care, social care, voluntary sector) | 232/363 | 63.9 |
| Worries about family getting COVID-19 infection | 227/363 | 62.5 |
| Lack of access to usual support from NHS mental health services | 196/362 | 54.1 |
| Increased risk from abusive domestic relationships | 192/360 | 53.3 |
| Increased difficulties for families/carers | 190/360 | 52.8 |
| Relapse and deterioration in mental health triggered by COVID-19 stresses | 182/363 | 50.1 |
| Increase in reliance on family/family tensions | 164/358 | 45.8 |
| High personal risk of severe consequences of COVID-19 infection (e.g. due to physical health comorbidities) | 155/359 | 43.2 |
| Difficulty engaging with remote appointments by phone or via digital platforms | 153/363 | 42.1 |
| Having to stay at home in poor circumstances, or not having a home to go to | 142/362 | 39.2 |
| Difficulty getting food, money or other basic resources | 138/362 | 38.1 |
| Diminished access to physical health care for problems other than COVID-19 | 131/362 | 36.2 |
| Difficulty understanding or following current government requirements on social distancing, self-isolation and/or shielding | 117/362 | 32.3 |
| Effects of COVID-19-related trauma | 113/360 | 31.4 |
| Risk of increased drug and alcohol use or gambling | 108/360 | 30.0 |
| Lack of access to or of equitable provision of physical healthcare for COVID-19 | 81/362 | 22.4 |
| Lack of access to medication and to processes for administering and monitoring it | 67/360 | 18.6 |
| Loss of liberty and rights due to changes in implementation of mental health legislation | 61/360 | 16.9 |
| Problems with police or other authorities because of lack of understanding of/ability to stick to current government requirements | 42/362 | 11.6 |

\*total respondents for this item

Similar problems were identified by the smaller group of 56 staff who reported working exclusively in perinatal services, although there were a few differences. Lack of access to usual support networks of family and friends was rated as very or extremely relevant by an even greater proportion (N=50/56; 89.3%), as was increased difficulties for families/carers (37/55; 67.3%). Worries about service users themselves getting COVID-19 (N=43/56; 76.8% rated very or extremely relevant) or their families getting COVID-19 (N=40/56; 71.4%) were also felt to be more relevant. Moreover, increased risk from abusive domestic relationships was identified as very or extremely relevant by 36/55 (65.5%): higher than in the larger sample.

Responses to free text questions revealed similar concerns. Staff were asked for which service users they were especially concerned and new mental health problems arising directly from the pandemic. The majority of staff reported being concerned about women in the perinatal period. Concerns and their impacts grouped as themes and how they inter-relate are displayed in Figure 1. Difficulties related either to the virus itself or environmental adaptations to reduce transmission. A few staff described cases of mental illness in women testing positive for COVID-19, including postpartum psychosis. Other staff reflected on the contribution of the current viral pandemic to the content of delusions arising in the context of new-onset or pre-existing psychosis. Another theme was that of fears of COVID-19 infection: *‘Pregnant mothers extremely worried about the risk of catching COVID-19 and the risks to their babies’*.

**Fig 1:**
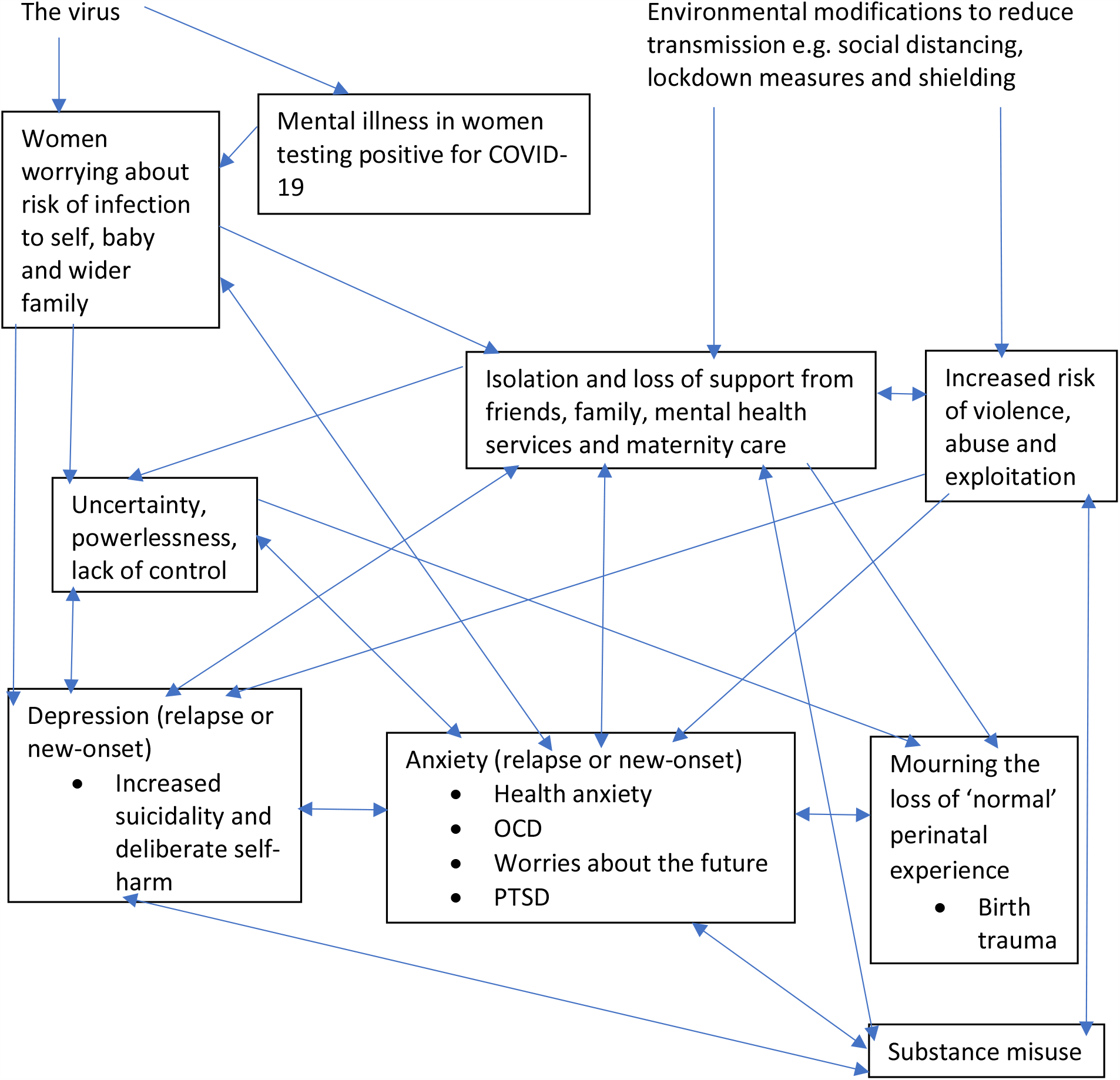
concerns from staff about areas of difficulty for perinatal service users and their perceived impacts (themes in boxes)

Other themes arose due to broader societal restrictions. Most staff reported concerns about loss of support and subsequent isolation in the context of reduced contact with family, friends and professionals during pregnancy and in the postpartum: *‘perinatal period … difficult time for some, involving some isolation. With the lockdown many pregnant women were left having remote appointments’*. Another closely related theme was concerns about those at an increased risk of domestic violence and abuse (DVA): *‘Vulnerable women and children at higher risk of DV’* and *‘Domestic abuse situations increasing’*, alongside feelings from staff of powerlessness to intervene, which will be considered further in the section that follows.

Most participants reported increases in depression and anxiety disorders among perinatal women, including worsening or relapse of pre-pandemic disorders or new symptoms since the pandemic’s onset. One staff respondent wrote: *‘Anxiety and OCD [Obsessive Compulsive Disorder] symptoms related to fear of infection’*. Another commented: *‘Impacting on mood and increasing risk of harm to self’*, when reflecting on the impact of social isolation. A few staff noted an increase in substance use: *‘increased substance misuse among perinatal women’*. Another theme which may have contributed to anxiety and depression in perinatal women was increased uncertainty and lack of control, already prominent during the perinatal period: *‘Feelings of being out of control’*. Finally, a few staff reflected that women’s experiences of having a baby during the pandemic had been very different to what they had expected: ‘*women due to give birth … very concerned about their birth experience’* and *‘Anxiety related to change in birth plans … Birth partner not being present at birth of baby’*. A possible increased risk of birth trauma was noted by some staff: *‘Birth trauma due to restrictions in maternity services’*. Many staff were particularly concerned about Post Traumatic Disorder (PTSD) in perinatal women in the longer-term.

### Changes to perinatal mental health care

Staff were also asked about changes made to services during the pandemic. 44.4% (92/207) of staff working in community teams estimated that referral rates declined by more than 10%. In inpatient services, 54.8% (17/31) of staff estimated that the monthly admission rate declined by more than 10%.

All respondents were asked to rate relevance of challenges to current work in the pandemic (since mid-March 2020). These differed somewhat between inpatient and residential settings (hospital inpatient service, crisis houses and residential services) and community settings (CMHTs, crisis teams and community groups). Four challenges rated most frequently as very or extremely relevant in inpatient and residential settings (N=58 staff) were: pressures of supporting colleagues (53.4% of respondents), adapting to new ways of working (53.4%), being infected with COVID-19 at work (51.7%) and putting infection control measures into practice (48.3%). The same challenges were identified in staff who reported working in MBUs specifically. In community settings, the most frequently rated as very or extremely relevant were: adapting to new ways of working (188/307; 61.2%), learning to use new technologies too quickly and/or without sufficient training and support (147/306; 48%), responding to additional mental health needs arising from COVID-19 (138/308; 44.8%) and pressures of supporting colleagues (123/305; 40.3%). The relative importance of these challenges across inpatient and community settings was broadly reflected in the smaller sample of staff who reported only working in perinatal settings. However, the proportion rating as very or extremely relevant ‘safeguarding and other risk management processes cannot be adequately mobilised due to limited social care, legal or police response’ was higher (51.8% in this sample versus 25.6% in the larger sample).

As all staff in the sample reported working with the perinatal population, they were later presented with additional challenges specific to perinatal work. Table 2 displays the proportion of staff rating each item as ‘very relevant’ or ‘extremely relevant’; items are ordered by highest proportion first. Completion rates of these items were just under 60% of the sample.

**Table 2:** challenges to perinatal work rated as very or extremely relevant by mental health staff.

|  | n/N* | % |
| --- | --- | --- |
| Challenges assessing mother and infant relationships because of lack of direct access | 90/208 | 43.3 |
| Difficulty planning and monitoring treatment due to reduced social care services | 67/209 | 32.1 |
| Difficulty planning and monitoring treatment due to reduced community midwife and health visitor services | 62/211 | 29.4 |
| Safeguarding procedures are more difficult than usual to mobilise | 62/211 | 29.4 |
| Reduced access to maternity units to carry out assessments | 59/211 | 28.0 |
| Referrals to our service not made or delayed because of the COVID-19 crisis | 58/210 | 27.6 |
| Reduced opportunities to admit to mother and baby units | 37/206 | 18.0 |
| Challenges arising from maternal or infant COVID-19 infection | 35/207 | 16.9 |
| Children are too readily taken into care because of obstacles to making other assessment and management plans at the present time | 9/208 | 4.3 |

\*total respondents for this item

Some challenges related to reduced ability to adequately assess and support perinatal women, their babies and their wider families, were echoed in responses to open questions. Themes of changes staff were most concerned about in the perinatal setting and their perceived impact are displayed in Figure 2. Community staff reported less frequent and fewer face-to-face contacts with women. Staff were concerned about their ability to detect early signs of mental illness when reviewing women remotely. Many were also worried about how to adequately assess and support the mother-infant interaction during the remote consultation by telephone or video: *‘unable to [do] vital work to help mums bond with their babies … only allowed to undertake phone calls not home visits*.*’* In extreme cases where staff are worried about the infant’s wellbeing and have safeguarding concerns, this assessment of interaction is part of the risk formulation as one staff respondent summarised: ‘*Vulnerable babies being cared for by unwell mothers who are not being identified due to lockdown because they are more isolated and less likely to be picked up by professionals’*. Risk assessment may also be challenging in a remote consultation: *‘Women with know[n] Domestic Abuse … doing video consultations so may not be able to be entirely honest about symptoms and risk’*. Another barrier to actioning safeguarding concerns identified by some staff was reduced provision from other agencies such as social services. Another theme of reduced community service provision in the postnatal period from other services, such as health visiting, was identified, which some staff felt impacted on breastfeeding support: ‘*not getting hands on support’*.

**Fig 2:**
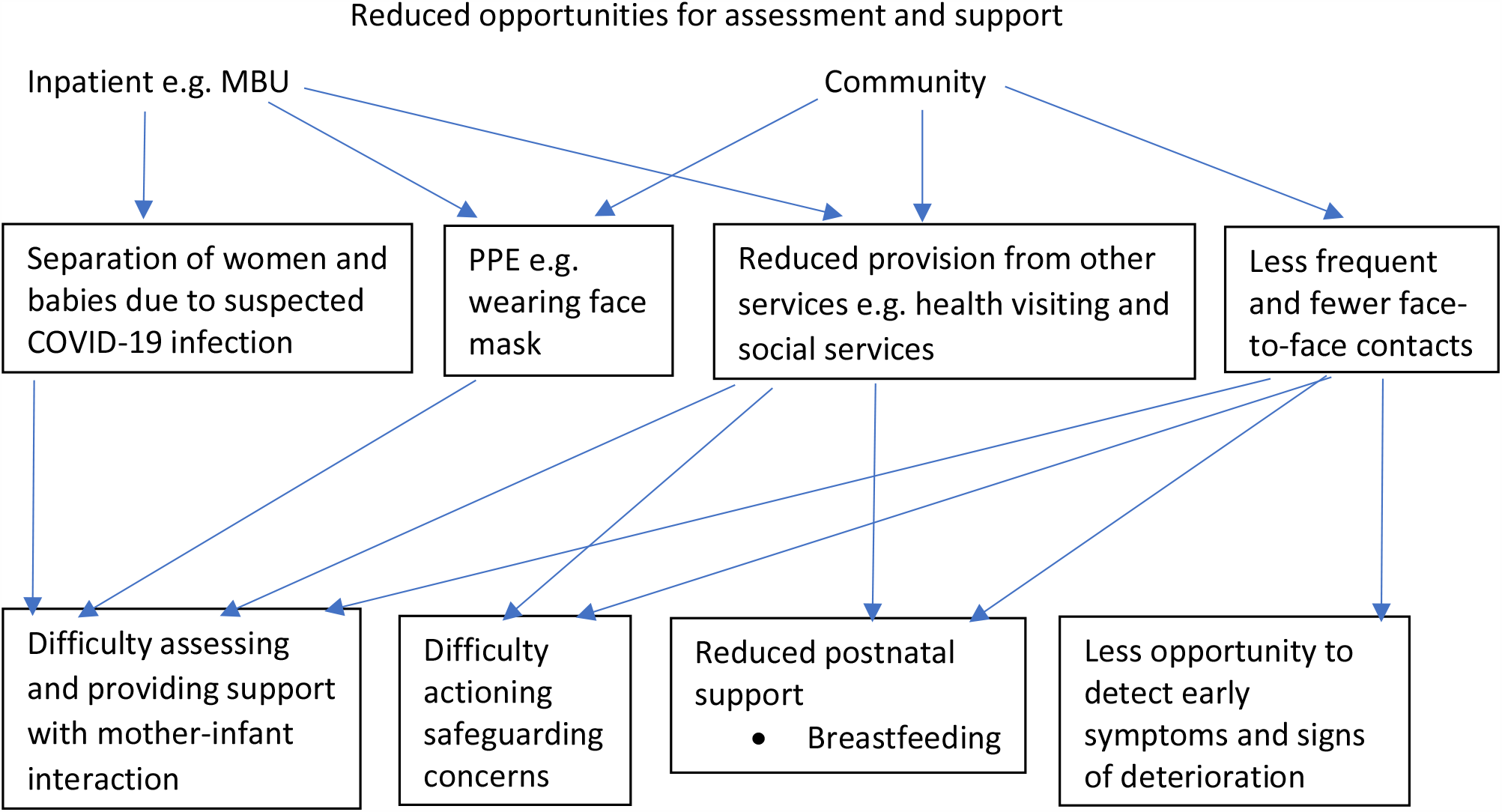
challenges to perinatal mental health service provision expressed by staff (themes in boxes)

Infection control measures presented some specific challenges in the inpatient perinatal setting to the assessment of the mother-infant interaction. A few staff working on MBUs reflected on the challenges posed by separation of women and babies with suspected COVID-19. Others were concerned about the impact of face mask wearing on the developing infant: *‘babies … cared for by masked nurses … lack of interaction with facial features*.*’*

Other open-ended questions elicited reflections on positive aspects of adaptations made to manage the impact which some staff felt would be beneficial to continue beyond the pandemic. Two themes of telemedicine benefits to service users and staff were identified. Several staff reflected on the flexibility of virtual appointments for women as busy new mothers and the range of online self-help and other supports available, such as cognitive behavioural therapy, mindfulness and third sector initiatives: ‘*local and national groups … stepping up with online support*.’ Benefits to staff were related to time management; reduced travel when working from home resulted in increased time for clinical record keeping and improved work life balance.

## Discussion

### Main findings

Mental health staff in this survey, administered during the COVID-19 pandemic lockdown in the UK, reported particular concerns about the mental health of women in the perinatal period compared with other service users. Perinatal women were perceived to be more vulnerable to the pandemic’s stressors, including social isolation, DVA and reduced service provision. While there has been little published research on the impact of the pandemic on perinatal mental health, pregnant women in China (Liu et al. 2020; Wu et al. 2020) and Canada (Saccone et al. 2020) assessed following, versus those assessed prior to, the onset of the pandemic, reported greater levels of anxiety and depression symptoms. However, to our knowledge, there has been no study to date reporting on staff perceptions of mental health care for perinatal women.

Staff working in the perinatal setting were concerned about how to support women during pregnancy and in the postpartum as a result of changes to care provided by their own and other services. Changes included reduced face-to-face contact in community mental health services, changes in infection control measures and reduced social care and home visiting services (i.e. maternity and health visiting services). Consequently, staff were worried about how to mobilise safeguarding procedures and assess and support the mother-infant interaction. There were also concerns about the impact of wearing masks on the developing infant; to our knowledge there has been no research investigating the impact of this in residential settings such as MBUs. However, staff highlighted positive changes such as the greater flexibility provided by remote consulting.

The impact of COVID-19 infection on women and their infants was inevitably a concern. Neuropsychiatric complications of infection have been reported, including acute confusion (Rogers et al. 2020), which may mimic postpartum psychosis.

### Strengths and limitations

This is the first study to document the experiences of mental health care staff supporting women in the perinatal period during the COVID-19 pandemic. The use of both qualitative and quantitative data facilitate triangulation of our findings (Tracy 2010).

However, there are some limitations posed by the convenience nature of the sample. Despite efforts to recruit a broad ethnic demographic, the number of non-White respondents in this sample is relatively small. There is also limited geographical diversity, with most of the respondents being from England; there may be different experiences in the UK’s devolved nations, with differing levels of perinatal mental health service provision, which were not adequately captured. Most of the staff were working in community services, with only 18 respondents from the MBU. There were also no questions about the experiences of partners of perinatal women so these experiences were not captured.

Furthermore, the sample included both staff working only in perinatal services and other staff working with perinatal women among other service users or in non-perinatal settings. Yet we believe the latter group is more representative of the structure of health services in many places outside the UK where there are no dedicated perinatal mental health services. Moreover, analyses in which only responses from staff working exclusively in perinatal services were analysed did not reveal substantial deviation from the larger sample.

### Implications and conclusions

As has been expressed by others (Holmes et al. 2020; Johnson et al. 2020) and highlighted in this study, some changes to service delivery will suit some groups of service users more than others so support must continue to be individually tailored. Indeed our study highlighted challenges to remote consulting unique to the perinatal period. Face to face assessment is necessary in high risk cases as highlighted by the recent confidential enquiry of maternal deaths in the UK during the first three months of the pandemic, which included four suicides and two domestic homicides (Knight et al. 2020).

Future research would usefully focus on how to manage these particular areas of difficulty such as the assessment of perinatal mental health, DVA and the mother-infant relationship (https://marcesociety.com/covid-19-perinatal-mental-health-resources/). Further research to elucidate the direct effect of COVID-19 infection on pregnant and breastfeeding women and the developing infant (Juan et al. 2020) would also help to address another major stressor for women during the pandemic.

In summary, there is an urgent need for research that provides an understanding of the experiences of perinatal women and their families during the COVID-19 pandemic. Greater understanding could illuminate potential targets for interventions by mental health staff and other professionals to support women with their perinatal mental health and safeguard women and their families against violence and abuse during the current pandemic and beyond.

### Lived experience commentary: Clare Dolman and Sarah Spring

This survey highlights the challenges encountered by mental health professionals trying to deliver vital services during the pandemic but reading their views on what they *thought* had affected women with perinatal mental illness the most during the pandemic, we wondered what women *themselves* would say had been most relevant to them. I understand that another survey is being conducted to answer those very questions, but perhaps that’s worth pointing out here.

There was no specific mention of the impact on fathers which one might expect when dealing with a perinatal population, though a couple of ‘difficulties’ related to families/carers. The results illustrate the detrimental effect of lack of face-to-face consultations, especially important to help women having problems with breastfeeding and bonding with their baby. Using video calls is referred to as a possible solution but they rely on a good connection, availability of the technology and a woman’s ability to use such technology if cognitively challenged when struggling with depression or other mental illness so reliance on this medium may exclude many mothers. For those women with developing psychosis and other conditions involving hallucinations, contact other than face-to-face could exacerbate or precipitate more delusions and, as mothers with these illnesses are good at disguising their symptoms, the severity of their illness could go undetected. This is another reason why remote contact should be used with extreme caution.

In relation to the service difficulties identified during the pandemic, it is noteworthy that perinatal services weren’t designated an essential service. This is an important point which should be considered in any future lockdown: many perinatal team staff were redeployed to hospital roles so women who had been considered serious enough to receive weekly therapeutic visits were only seen much less frequently thus increasing the risk that their condition would worsen. Similarly numbers of Health Visitors, who provide essential support to women in the perinatal period, were redeployed. This policy now appears short-sighted and in need of review.

This is an independently written perspective from lived experience contributed by some of the members of the IOPPN’s Women’s Mental Health Section’s service user perinatal advisory group with relevant experience.

## Data Availability

The survey dataset is currently being used for additional research by the author research group and is not currently available in a data repository. A copy of the survey is available at this web address: https://opinio.ucl.ac.uk/s?s=67819.

## Authors’ contributions

SJ drafted the survey and CDL developed the online version. CDL and AS led on recruitment strategies. SJ and LMH conceived the idea for the perinatal study. CDL cleaned the data. CAW designed the study analysis plan and analysed the data. Peer debriefing of qualitative data was provided by LMH to CAW. CAW wrote the first draft of the manuscript with input from LMH. All authors critically revised this draft and have approved the final version.

## Acknowledgements

We thank the clinicians who took the time during the pressures of the pandemic to complete the survey, including those who provided feedback on a pilot survey.

## Declarations

### Ethics approval

The King’s College London research ethics committee approved this study (MRA-19/20-18372).

### Consent to participate

Information on participation was provided on the front page of the survey. By starting the survey, participants agreed that they had read and understood all of this information.

### Consent for publication

It was explained on the front page of the survey that responses may be used in articles published in scientific journals and that these articles would not include any information which could be used to identify any participant.

### Code availability

Not applicable.

